# A Role For Electrocardiograms in Aortic Stenosis Screening: A Retrospective Trial

**DOI:** 10.1101/2024.04.10.24305584

**Authors:** Matthew Pulaski, Aditi Newadkar, Iman Richie, Cole Urnes, Ivan Ho, Jubin Joseph, Ray Matthews, Junaid Zaman

**Affiliations:** Keck School of Medicine

**Author notes:** **Corresponding author:** Matthew Pulaski, MD, (732)757-8559. **Mailing address of all authors:** Keck School of Medicine, 2020 Zonal Avenue, Los Angeles CA 90033.

**Keywords:** Aortic stenosis, screening, cardiology

## Abstract

**Background:** Aortic stenosis affects 1 in 50 adults over age 65 and is associated with significant morbidity and mortality. Machine learning has identified an association between right-sided precordial U waves and moderate to severe aortic stenosis. No study has explored the role of ECG screening by primary care physicians for patients with unknown aortic stenosis status.

**Methods:** A retrospective single center cohort analysis performed by non-cardiologists identified right-sided precordial U waves on ECGs in fifty adults ages 65 to 89. Following identification, reviewers were unblinded to echocardiograms to determine whether there was an association between right-sided precordial U waves and aortic stenosis severity. Fifty age- and gender-matched patients without right-sided precordial U waves comprised the control group.

**Results:** Chi-squared analysis revealed a significant association between right-sided precordial U waves and severity of aortic stenosis (χ^2^ = 16.77, df = 3, p < 0.001). Multinomial logistic regressions demonstrated no relationship between categorical SBP (< 125, 126 – 145, > 145) and aortic stenosis (p = 0.35), but increasing categorical age (65-73, 74-81, 82-89) was associated with moderate to severe aortic stenosis (p = 0.002).

**Conclusions:** Our data highlights right-sided precordial U wave identification by non-cardiologists as a novel objective adjunct to the physical examination in detection of a medical condition which portends significant morbidity and mortality if left untreated. These findings motivate a prospective randomized clinical trial in the utility of right-sided precordial U waves in screening for aortic stenosis in primary care settings.

## Introduction

Aortic stenosis (AS) is the most common valvular heart disease in developed countries, affecting 1 in 50 adults over age 65 and portending significant morbidity and mortality.^1^ The majority of patients with AS are asymptomatic, and detection of AS by auscultation has been shown in recent meta-analysis to be highly variable^2^, hence adjunctive methods are needed. Recently, machine learning has identified an association between right-sided precordial U waves (RSPUW) on electrocardiograms (ECG) and moderate to severe aortic stenosis.^3,4^ Among patients with known severe AS undergoing transcatheter aortic valve replacement (TAVR), 62% were found to have U waves on pre-TAVR ECG with 87% of those exhibiting U-wave diminution on post-TAVR ECG.^5^ To our knowledge, no study has explored the role of ECG screening in patients with unknown AS status. We therefore investigated whether general practitioners blinded to AS disease status are able to identify RSPUW on ECGs, and whether RSPUW are correlated with AS severity.

## Methods

A retrospective single center cohort analysis was approved by the University of Southern California institutional review board and performed by non-cardiologists (internal medicine residents and internists). Inclusion criteria were age 65-89, diagnosis of hypertension (HTN), prior ECG and echocardiogram. Exclusion criteria were atrial fibrillation or flutter, ventricular pacing, left bundle branch block, bradycardia (heart rate < 50), hypokalemia (K < 3), or prior AVR.

ECGs with RSPUW in consecutive leads (at least two of V1-V3), with ≥ 2 U waves per lead with amplitude ≥ 0.5 mm, formed the U+ cohort. Reviewers were then unblinded to the echocardiogram obtained within one year of the ECG to assess for presence and severity of AS, graded by American Society of Echocardiography guidelines. Retrospective data collection was continued until n= 50 patients were obtained in both U+ and U-cohorts, matched for age and gender.

Chi-squared analyses assessed the relationship between U+ status and AS severity (none, mild, moderate, severe). We used multinomial logistic regression to investigate the relationship between age and systolic blood pressure (SBP) on moderate to severe AS.

## Results

In total, 976 and 294 patient charts were reviewed to identify 50 patients for the U+ and U-cohorts, respectively. See Table 1 for demographic data. Median time between ECG and lab collection (to ensure a K ≥ 3) was 0 days for both cohorts. Median time between ECG and echocardiogram was 1 day and 1.5 days respectively for U+ and U-arms. Based on a chi-square test of independence, there is no statistically significant difference in racial/ethnic demographics between U+ and U-cohorts (χ^2^ = 3.63, df =4, p = 0.46). Chi-squared analysis revealed a significant association between U+ status and severity of aortic stenosis (χ^2^ = 16.77, df = 3, p < 0.001). Multinomial logistic regressions demonstrated no relationship between categorical SBP (< 125, 126 – 145, > 145) and AS (p = 0.35), but increasing categorical age (65-73, 74-81, 82-89) was associated with moderate to severe AS (p = 0.002).

**Table 1:** Demographic, Echocardiogram and ECG Data with Unpaired T-Tests.

|  | U+ Cohort (95% CI) | U-Cohort (95% CI) | p value |
| --- | --- | --- | --- |
| **Demographic Data** |  |  |  |
| Age | 75.6 (73.7-77.5) | 75.6 (73.7-77.5) | 1.00 |
| Gender | 26/50 M (52%) | 25/50 M (50%) | N/A |
| Body Surface Area, m<sup>2</sup> | 1.9 (1.8-1.9) | 1.9 (1.8-2.0) | .60 |
| Non-Hispanic White | 29 (58%) | 23 (46%) | - |
| Non-Hispanic Black | 2 (4%) | 1 (2%) | - |
| Hispanic | 14 (28%) | 15 (30%) | - |
| Asian/Pacific Islander | 4 (8%) | 10 (20%) | - |
| Other Race/Ethnicity (Not Listed) | 1 (2%) | 1 (2%) | - |
| **Echocardiogram Data** |  |  |  |
| BP systolic, mmHg | 136 (130-142) | 125 (120-130) | **.03** |
| BP diastolic, mmHg | 66 (63-70) | 65 (62-68) | .53 |
| LVEF, % | 60 (58-63) | 58 (54-62) | .39 |
| IVSd, cm | 1.2 (1.1-1.3) | 1.1 (1.0-1.2) | .30 |
| LVPWd, cm | 1.1 (1.0-1.2) | 1.0 (1.0-1.1) | .23 |
| Peak Velocity, m/s | 2.5 (2.2-2.9) | 1.7 (1.5-2.0) | **<.001** |
| Mean Gradient, mmHg | 27 (22-33) | 13 (9-16) | **.01** |
| AVA, cm<sup>2</sup> | 1.5 (1.3-1.8) | 2.0 (1.8-2.3) | **.01** |

| **ECG Data** |  |  |  |
| --- | --- | --- | --- |
| U wave Amplitude, mm | 0.79 (0.71-0.86) | 0.05 (0.02-0.08) | **<.001** |
| **Aortic Stenosis Data** |  |  |  |
| Moderate or severe AS | 18 (36%) | 4 (8%) | **.001** |

## Discussion

Our findings support the use of RSPUW, evaluated by non-cardiologists, in detection of moderate to severe AS. ECGs are inexpensive and routine tests performed at annual physical examinations in the older population where AS occurs. Our data highlight the RSPUW as a novel objective adjunct to the physical examination in detection of a medical condition which portends significant morbidity and mortality if left untreated. These findings motivate a prospective randomized clinical trial in the utility of RSPUW in screening for AS in primary care settings.

This was a small retrospective study at an academic medical center with high TAVR volume. Hence the findings need to be replicated in non-specialist settings. Additionally, U wave reading was manual. To our knowledge, no ECG artificial intelligence software offers U wave analytics, and collaborative efforts are needed to fully automate RSPUW detection in order to be truly objective. Echocardiograms were included within 1 year, but more contemporaneous data will be needed in a prospective trial.

## Data Availability

Data is maintained on a private, external server and will not be made publicly available with the publication of the study.

## Conflict of Interest Statement

There are no conflicts of interest to report for all authors.

## Notes

### Competing Interest Statement

The authors have declared no competing interest.

### Funding Statement

No funding was accessed or obtained in the execution of this study.

### Author Declarations

Ethics committee/IRB of University of Southern California gave ethical approval for this work

## References

1. Nathaniel S, Saligram S, Innasimuthu AL. Aortic stenosis: An update. World J Cardiol. 2010;2(6):135–139. doi:10.4330/wjc.v2.i6.135

2. Davidsen AH, Andersen S, Halvorsen PA, Schirmer H, Reierth E, Melbye H. Diagnostic accuracy of heart auscultation for detecting valve disease: a systematic review. BMJ Open. 2023;13(3):e068121. doi:10.1136/bmjopen-2022-068121

3. Kwon J, Lee SY, Jeon K, et al. Deep Learning–Based Algorithm for Detecting Aortic Stenosis Using Electrocardiography. J Am Heart Assoc. 2020;9(7):e014717. doi:10.1161/JAHA.119.014717

4. Johnson K, Torres Soto J, Glicksberg B, et al. Artificial Intelligence in Cardiology. J Am Coll Cardiol. 2018 Jun, 71 (23) 2668–2679. 10.1016/j.jacc.2018.03.521

5. Lampert J, Pulaski M, Miller MA, et al. Right Precordial U Waves in Severe Aortic Stenosis: “Real-World” Prevalence and Diminution by TAVR. J Am Coll Cardiol. 2022;79(24):2467–2469. doi:10.1016/j.jacc.2022.04.012

